# A Time Series Analysis and Predictive Modeling of COVID-19 Impacts in the African American Community

**DOI:** 10.1101/2021.05.13.21257189

**Authors:** Timothy Oladunni, Sourou Tossou, Max Denis, Esther Ososanya, Joseph Adesina

## Abstract

**Background:** Sometimes in 2019, there was an outbreak of coronavirus pandemic. Data shows that the virus has infected millions of people and claimed thousands of lives. Vaccination and other non-pharmacological interventions have brought a relief; however, COVID-19 left some indelible marks. This work focuses on a time series analysis and prediction of COVID-19 fatality rates in the Black community. Decision makers will find the work useful in building a robust architecture for a resilient pandemic preparedness and responsiveness against the next pandemic. Method: Our analysis of COVID-19 cases and deaths spans March 2020 to December 2020. Assuming there was no vaccine and other factors remained the same, we hypothesized that COVID-19 disproportionality would have continued. To test our hypothesis, COVID-19 forecasting cases and deaths models were built for the total population as well as the Black population. Holt and Holt-Winters exponential smoothing forecast methodologies were used for the forecast modeling. Forecasting accuracy was based on Mean Absolute Percentage Error (MAPE). Furthermore, we designed, developed, and evaluated a fatality rate predictive model for a Black county. Considering the number of ethnic groups in the USA, a Black county was defined as any county in the USA that at least 45% of its population are Blacks. Five learning algorithms were trained and evaluated. Dataset was a merger of datasets obtained from John Hopkins COVID-19 repository, US Census Bureau and US Center for Disease Control and Prevention.

**Results and Conclusion:** Time series analysis shows that there exists a strong evidence of COVID-19 disproportionate impacts in the states investigated. Using 9 different criteria for performance comparison, our predictive modeling showed that decision tree model has a slight edge over other models. Our experiment suggests that Blacks and senior citizens with pre-existing condition living in Georgia State are the most vulnerable to COVID-19.

## I. INTRODUCTION

In the United States, preliminary statistical data shows that the Black population were disproportionally affected by the pandemic (Laurencin & McClinton, 2020). Time series analysis points to the fact that the pandemic had a devastating, disruptive, and damaging impact on the Black community (Fletcher, et al., 2020). Since the world woke up to the emergence of this diseases in the late 2019 (Morens, et al., 2020), fatalities and mortalities rates for the most part were on an upward trajectory among this community (R.B.Hawkins, E.J.Charles, & J.H.Mehaffeyab, 2020). Counties and zip codes with large population of Blacks became synonymous with high COVID-19 fatalities. For example, Blacks in Prince Georges, Montgomery, and Baltimore counties are 32%, 20.1% and 30.3% respectively of the total population of the state of Maryland in the United. As of January 7, 2021, these three counties were first, second and third respectively in the number of coronavirus cases in the state. On the fatalities count, Montgomery county displaced Prince Georges county for the top spot. Prince Georges county came second while Baltimore county retained the third position (New York Times, 2021).

Scientific evidence has connected the disproportionate impact of coronavirus cases in the Black community to the poor health care and social economic disadvantage of the community (Tai, Shah, Doubeni, Sia, & Mark L Wieland, 2020). Before the outbreak of the COVID-19 pandemic, researchers found that the life expectancy of the Blacks are lower than other ethnic groups. Study shows that the average life expectancy of White and Black women are 81.0 years and 78.1 years respectively. Furthermore, average life expectancy of White male to the Black male are found to be 76.1 year and 71.5 years respectively (M., 1985). The disproportionality in the life expectancy of Blacks when compared with the whites are not surprising. This is because several studies have shown that African Americans have been bearing the burden of most diseases in the US. For example, research on diabetics shows that Blacks have more than 60% chance of being diagnosed with the disease than the Whites (U.S. Department of Health and Human Services Office of Minority Health. Diabetes and African Americans). They also have 42% chance of being a new victim of HIV (Cneters for Disease Control and Prevention). Furthermore, heart attacks data shows that the population are 20% more susceptible than the White (U.S. Department of Health and Human Services Office of Minority Health. Heart Disease and African Americans). The same narrative has been found to be true for obesity (U.S. Department of Health and Human Services Office of Minority Health. Obesity and African Americans) and asthma (Centers for Disease Control and Prevention). Several studies have also shown that African Americans are more likely to die prematurely from any diseases than Whites (Cunningham, et al., 2017).

The prevalent poor health care system coupled with long standing pre-existing conditions have been found to have a drastic, significant, and far-reaching impact on the mortality and fatality rates of African American COVID-19 patients (Richardson, Hirsch, & Narasimhan, 2020). Patients with diseases such as hypertension, diabetes, congestive heart failure, chronic kidney disease and cancer have been found to have a higher mortality and fatality rates when contracted COVID-19 (Ssentongo, Ssentongo, Heilbrunn, Ba, & Chinchilli, 2020). Ferdinand and Nasser argued that the prevalent cardiovascular disease (CVD) among African Americans which directly links with a poor health care condition of the community is to be blamed for the disproportionality in the coronavirus cases in the African American and other minority communities (Ferdinand & Samar A. Nasser, 2020). Socio-economic factors have also been found to contribute to the disproportionality in the coronavirus cases in the African American community. Studies have shown that Blacks and Whites have a poverty rate of 22% and 9% respectively (State Health Facts, 2019). Furthermore, the median household income of the Whites is found to be 10 times that of the Blacks (Kochar & Cilluffo, 2017). During the pandemic, only 20% of African American population could work from home as compared with the 30% of Whites. Furthermore, 34% and 14% of African Americans and whites are likely to use public transportations respectively (Anderson, 2016). Unhealthy diet (Dutko, Ploeg, & Farrigan, August 2012) and population density (Branigin, 2020) are other contributing factors.

Since the outbreak of the coronavirus pandemic, there have been different studies on COVID-19. However, there seems to be a gap in literature on a time series analysis and predictive modeling of fatality rates in the African American community. The vulnerability of the community to coronavirus pandemic has been a subject of different studies (Dyer, 2020). We believe that all stakeholders should have an answer to the question; is there COVID-19 healthcare disproportionality in the African American community? If the answer is yes, the next question is: how do we predict COVID-19 fatality rate in the African American community? Such an answer will help in improving the quality of cares to the affected population. Therefore, using scientific principle, we analyzed COVID 19 datasets from April 12, 2020 to December 25, 2020; to understand hidden patterns and discover knowledge on the disproportionate impact of the coronavirus pandemic and predict fatality rate in the disadvantaged demography (Dowd, et al., 2020).

The spread of the coronavirus was controlled in parts with measures such as border closing, travel restriction and airport screening. However, research shows that these measures only reduced the spread of the disease without a far-reaching effect on its impact (Wells, et al., 2020). To improve mitigation strategy, most governments encouraged handwashing (Shigute, Mebratie, Alemu, & Bedi, 2020), social distancing (Andersen, 2020) and mask covering (Rab, Javaid, & Haleem, 2020). We argue that a disproportionate impact of a pandemic is a major setback to an effective universal mitigation strategy.

In this paper, understanding COVID-19 health care disproportionate impact in the Black community is our goal. Achieving this goal, we will consider the following 3 objectives, 1) COVID-19 Time Series Analysis, 2) COVID-19 Health Care Disproportionality and African American Community, and 3) Predicting COVID-19 Fatality Rate in African American Community. The paper is organized as follows; COVID-19 times series analysis, COVID-19 healthcare disproportionality and Predicting COVID-19 Fatality Rate in African American Community are discussed in sections 2, 3, and 4 respectively. We conclude our study in section 5 with the implication of the study in section 6. We highlight the limitation of the study, future work and acknowledge our funding source in sections 7, 8 and 9 respectively.

## 2. COVID-19 TIME SERIES ANALYSIS

Dataset for this experiment was obtained from The COVID Racial Data Tracker. The repository is a collaboration project between the COVID Tracking Project and Boston University Center for Antiracist Research (Atlantic, 2020). COVID 19 datasets from April 12, 2020 to December 25, 2020 was extracted for the states of Floria (FL), Georgia (GA), Maryland (MD), Mississippi (MS), North Carolina (NC), Philadelphia (PA), South Carolina (SC) and Virgina (VA). These states have medium to large population of African Americans in the USA. We began our investigation by exploring the dataset to see underlying patterns, trends and seasonalities at different time lines.

We ploted area graphs to show the visual representation of the underlining patterns of the dataset. Graphical representation was shown for the total cases and deaths in each of the states. Cases and deaths of Blacks were also plotted for each state. Research has shown that graphical data visualization has a direct impact on the effectiveness of data analysis. Thus visual methodology of data exploration and representation has a unique way of making information noticeable, salient and memorable (Zacks, Levy, Tversky, & Schiano, 2002). This suggests that visual expressions improves communication. An area graph combines the attributes of line and bar charts. In our analysis, states are represented with shaded areas staked on the top of one another. This arrangement shows how the impact of the virus changes over time in each of the states. Our graph is broken down further into yearly quarters. At the end of each quarter, we show the state of the virus in each of the states. Figures 1, 2, 3 and 4 show the area graphs of total cases, Black cases, total death and Black deaths respectively.

**Figure 1.**
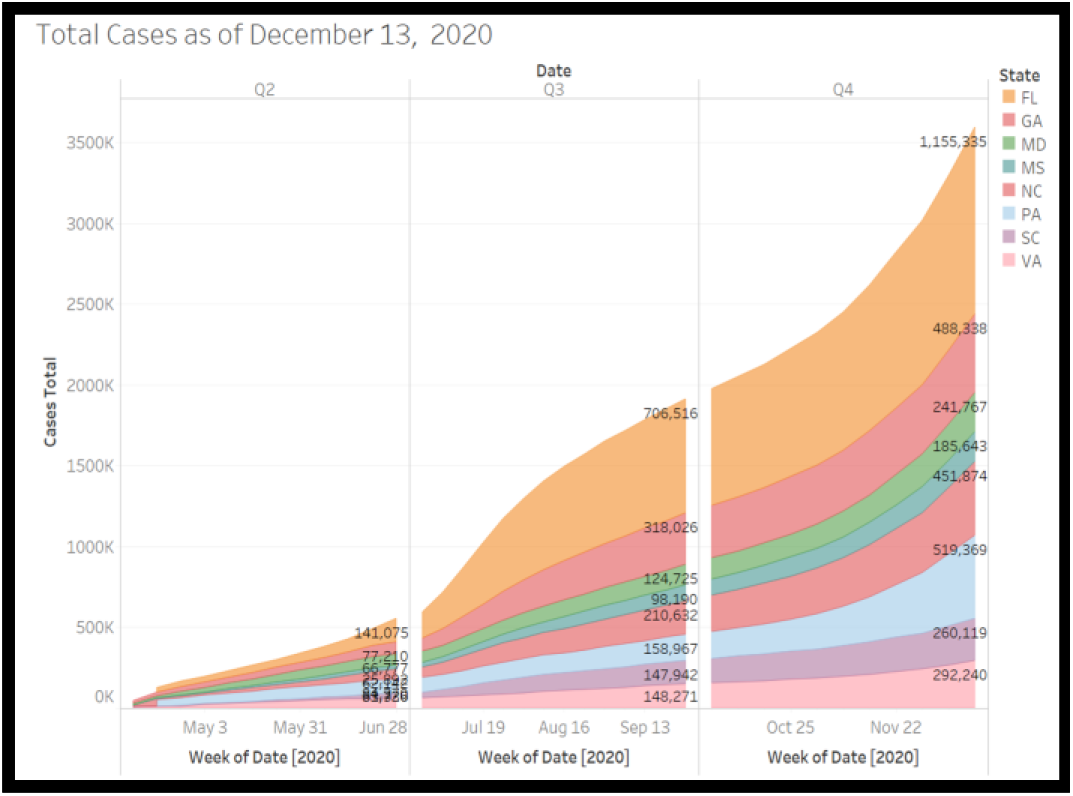
Area graph of the Total COVID 19 cases in FL, GA, MD, NC, SC, MS, PA and VA

**Figure 2.**
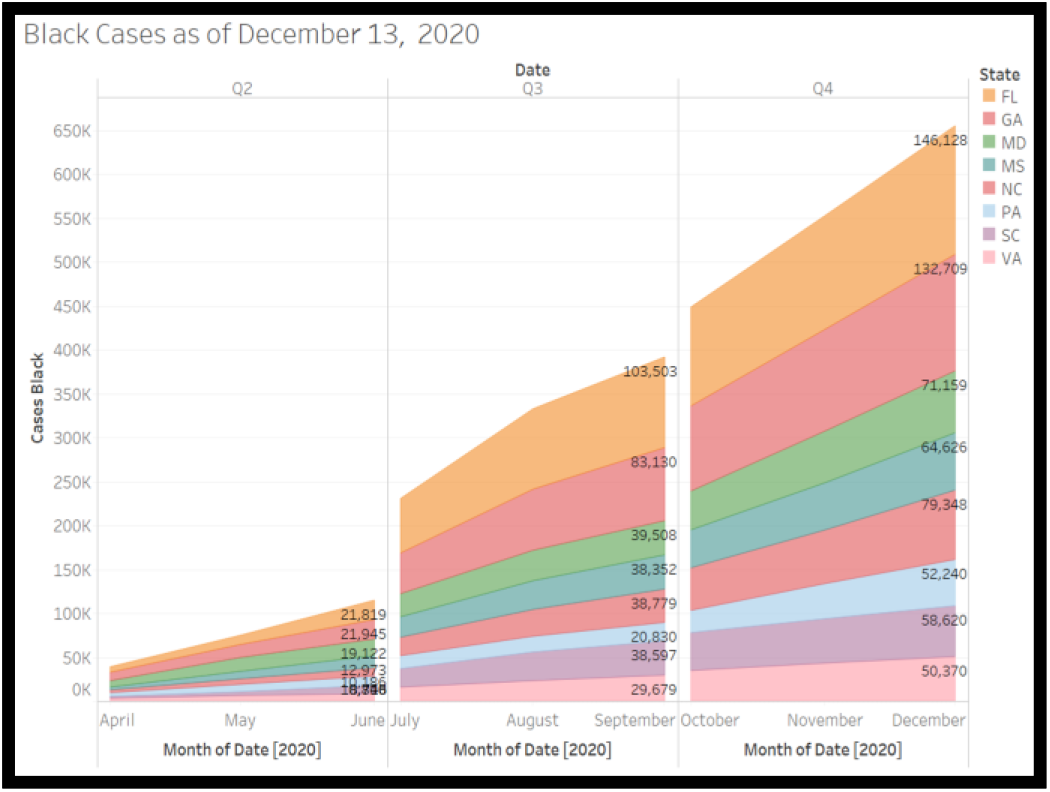
Area graph of Black COVID 19 cases in FL, GA, MD, NC, SC, MS, PA and VA

**Figure 3.**
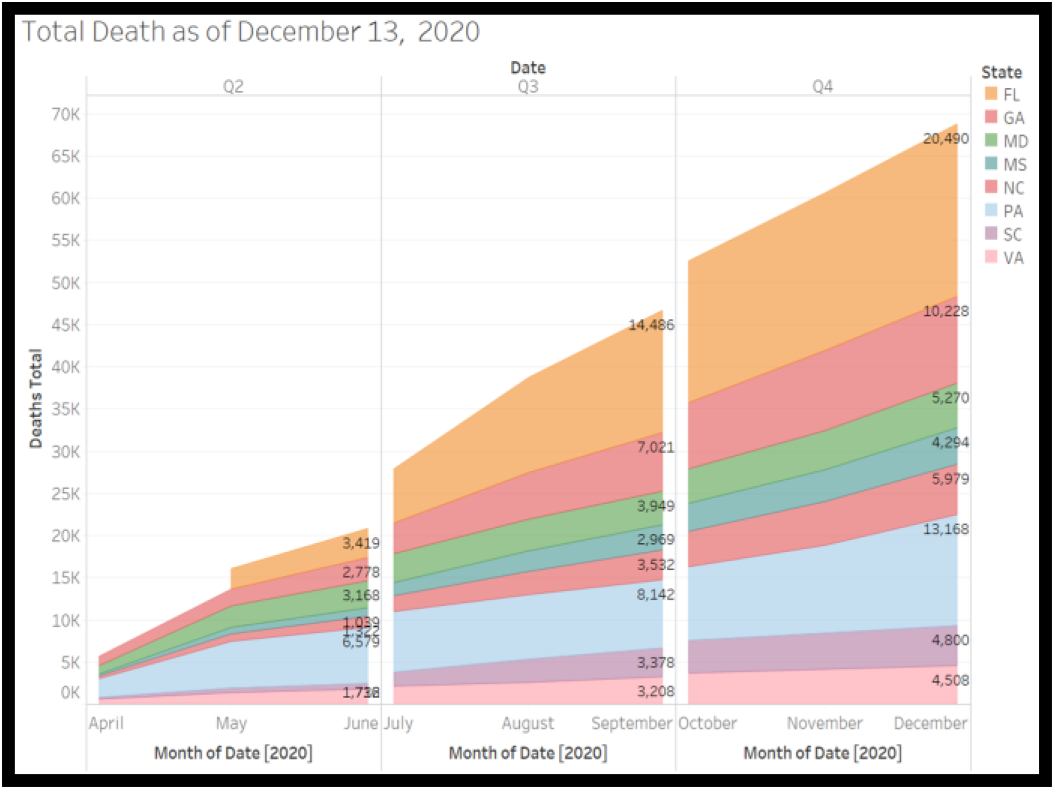
Area graph of the Total COVID 19 deaths in FL, GA, MD, NC, SC, MS, PA and VA

**Figure 4.**
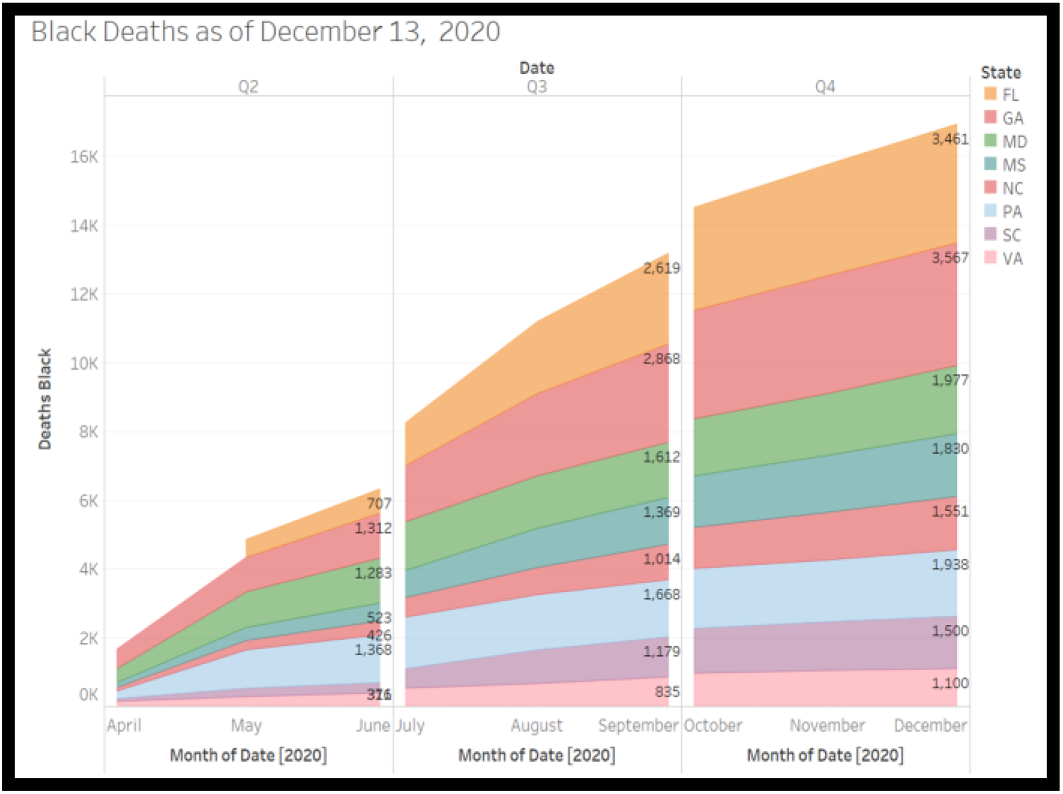
Area graph of the Black COVID 19 deaths in FL, GA, MD, NC, SC, MS, PA and VA

As shown in figure 1, at the end of each quarter, many states have seen their share of the affliction of COVID-19 followed different trajectories. For example, at the end of the first quarter, FL was approximately one hundred and forty thousand total cases. However at the end of the second quarter, cases at FL has skyrocketed to over seven hundred thousand. This is a five time increment. As shown in the graph, the virus timeline was divided into 3 quarters. This is because the pandemic impacted the United States sometimes in March 2020 (Centers for Disease Control and Prevention). Although it had began in China in 2019 (nature) and the World Health Organization declared it a pandemic in March 11, 2020 (WHO Director-General’s opening remarks at the media briefing on COVID-19 - 11 March 2020).

The trajectory of the graph shows that the first quarter of COVID-19 started at the begining of April. Therefore, April to June is COVID-19 first quarter. As shown in the graphs, at the first quarter, the virus had a little impact in all the states. The major impact of the virus became very abovious at the second quarter (July to September). The virus slowed down at the early second quarter but gained momentum towards the end of the quarter. The brief period when the virus slowed down might be the beginning of the summer period in the US. However, a further looks at the graph shows that the high temperature assumption of the summer period did not last. By August, the situation at FL was alarming, the virus seemed to have a found an abode in the Sunshine state.

As the total number of COVID-19 cases were rising in each of these states, fatalitity rate was on increase. Figure 3 shows that at the end of the first quarter, PA was hardest hit with coronavirus deaths. The state recorded approximatley 6 500 deaths. COVID-19 victims at NC was a liitle above 1 000, FL was at 3 400. However at end of the third quarter, FL took the lead with more than 14 000 deaths (this is around 300% increase). It seems PA been able to bend the curve; its death count increased to only 8 000 (this is around 23% increase).

Except for GA which was close to FL in high death counts, other states closed the second quarters with approximately 3 000 COVID-19 victims. By December 16, FL, PA and GA lead the death tolls and recorded more than 20 thousands, 13 thousands and 10 thousands deaths respectively. Other states were at around 4 000 and 5 000.

## 3. COVID-19 HEALTH CARE DISPROPORTIONALITY (CHCD) AND AFRICAN AMERICAN COMMUNITY

The time series analysis shows the underlying patterns of the waves of COVID-19 in the states under investigation. Figures 1 to 4 suggest that there is a disproportionality in the percentage of Blacks who contracted COVID-19 to the percentage who died of it. For example, as of December 13, 2020, in FL, the total cases and Black cases were 1 155 335 and 146 128 respectively-this is a 12.65% of Black cases to the total cases. However, the total COVID-19 deaths and Black deaths were 20 490 and 3 461 respectively-which is a 16.89% of Black deaths to the total deaths. Also, in GA there were total cases and Black cases of 488 338 and 132 709 respectively-this is a 27.18% Black cases to the total cases. However, there were total deaths and Black deaths of 10 228 and 3 567 respectively-this is 34.87% Black deaths to the total deaths. In both FL and GA, the results suggest a disproportionality of 4.24% and 7.70% respectively. Table 1 was created for all the eight states in our study.

**Table 1.** COVID-19 Health Care Disproportionality

| State | Total Cases | Black Cases | (Black Cases/Total Cases) % | Total Death | Black Death | (Black Death/Total Death) % | CHCD |
| --- | --- | --- | --- | --- | --- | --- | --- |
| FL | 1155335 | 146128 | 12.65% | 20490 | 3461 | 16.89% | 4.24% |
| GA | 488338 | 132709 | 27.18% | 10228 | 3567 | 34.87% | 7.70% |
| MD | 241767 | 71159 | 29.43% | 5270 | 1977 | 37.51% | 8.08% |
| MS | 185643 | 64626 | 34.81% | 4294 | 1830 | 42.62% | 7.81% |
| NC | 451874 | 79348 | 17.56% | 5979 | 1551 | 25.94% | 8.38% |
| PA | 519369 | 52240 | 10.06% | 13168 | 1938 | 14.72% | 4.66% |
| SC | 260119 | 58620 | 22.54% | 4800 | 1500 | 31.25% | 8.71% |
| VA | 292240 | 50370 | 17.24% | 4508 | 1100 | 24.40% | 7.17% |

We define COVID-19 Health Care Disproportionality (CHCD) as the difference between the percentage of Black Cases to Total Cases and Black Deaths to Total Deaths. If the result of the former is more than the later, it suggests that there is a disproportionality in COVID-19 impacts.

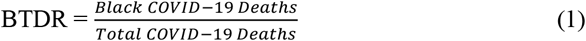

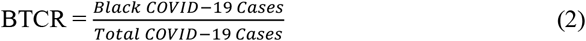

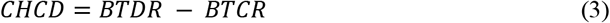

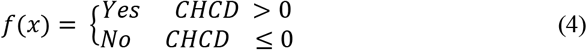

Table 1 shows the result of CHCD as of December 13, 2020. The table suggests that MD, NC and SC are doing worse than other states in the proportion of Blacks who survived COVID-19 when compared with the number of those who contracted it.

### Forecasting CHCD

Our objective in this section is to demonstrate that CHCD will continue in the Black community. Achieving this objective, we will: 1) build models with the capability of forecasting COVID-19 total cases and number of Blacks who will likely contrast COVID 19, 2) build models to forecast COVID-19 total deaths and Black deaths, and 3) compute a COVID 19 Health Care Disproportionality Table. *Suppose there is no vaccination and other factors remain the same, we hypothesized that CHCD will continue in the Black community*.

We designed, developed, and evaluated 8 forecasting models. The proposed models will forecast total cases, total death, Black cases and Black deaths using Holts and Holt-Winter exponential smoothing. An exponential smoothing forecast is a univariate time series forecast methodology. Its uniqueness is on its exponential decaying average of the weights of past observations; most recent observations are apportioned more weights than old observations. This approach makes it more reliable and accurate in forecasting wider range of time series than the moving average. Our forecast will be to the end of 2021 first quarter.

We experimented to determine CHCD for the first quarter of 2021 in the states under investigation. As shown in figure 5, dataset went through a pre-processing stage to make it suitable for the experiment. At the design and development stage, forecasting models were designed and developed using Holts and Holts-Winters methodologies. We evaluated the performance of each models at the evaluation stage. Selection of best models was done at the select best models’ stage. Computation of Black/Total Death Ratio (BTDR) and Black/Total Case Ratio (BTCR) was done at the compute BTDR and BTCR respectively. Finally, a CHDR table was created at the CHDR table stage.

**Figure 5.**
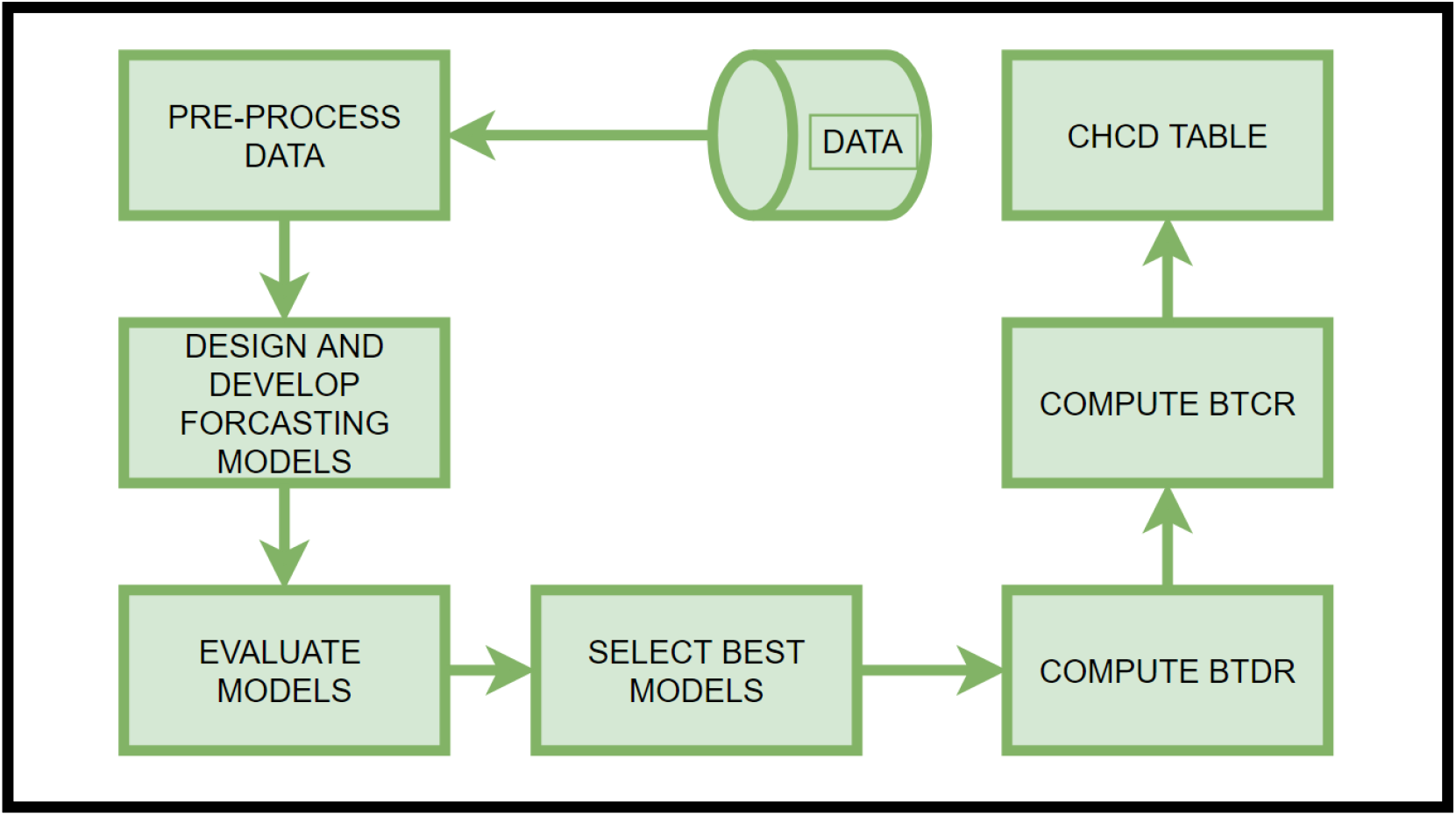
The Architectural Layout of the Experiment

#### 3.1.1 Simple Exponential Smoothing

The simple exponential smoothing does not consider trend or seasonality in forecast modeling. Therefore, it is not a good estimator of a time series where trend or seasonality is a factor. This puts a limit to the effectiveness of its application. In the simplest form, the forecasted value *y*′_*t+h*|*T*_ is equal to the last observed value *y*_*T*_ for the step ahead *h = 1, 2, …. N*

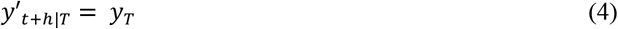

Equation 4 can be simplified further to be weighted average of all past observations.

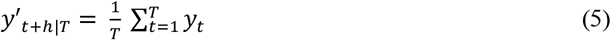

Equation 5 suggests that all observations are equally important; equal weights are assigned to each of them. We can improve on this assumption by including decaying weights to past observations (most recent observations are assigned largest weights, while oldest observations are assigned smallest weights). This is the intuition of exponential smoothening:

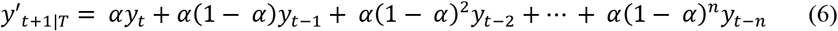

y_1_, …., y_t_ are t observations. The decay rate is represented as parameter *α*; where 0 ≤ *α* ≤ 1. As *α* moves towards 1, the most past observations are given more weights; making the learning rate to become faster. On the other hand, a value close to 0 reduces the learning rate because more weights are given to the past observations.

#### 3.1.2 Holt (Double Exponential Smoothing)

Holt exponential forecast is an extension of the simple exponential forecast methodology. It includes the trend smoothing parameter *β* for the trend *b*_*t*_ in addition to the decay rate *α* for the smoothing parameter at level *l*_*t*_ of a time series *t*. This improves the effectiveness and accuracy of its forecasting capability.

The estimated forecast *y*′_*t+h*|*t*_ consist of the level *l*_*t*_ *and trend b*_*t*_ for steps ahead *h = 1, 2, …. n*, of the series at time t.

The level *l*_*t*_ can be expressed as

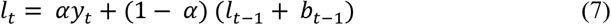

The Trend *b*_*t*_ can also be expressed as

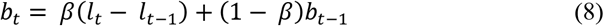

Combining equations 7 and 8, the Holt Forecasting equation is defined as

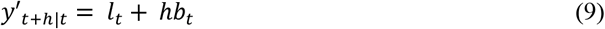

Equation 9 suggests that the forecasting equation is no longer flat. As shown, the h-step ahead forecast now depends on the estimated levels as well as the trend estimate of a time series *t*. The major short comings of the double exponential smoothing is its incapability to account for seasonality in time series forecasting.

#### 3.3.4 Holts-Winter (Triple Exponential Smoothing)

The Holts-Winter is an improvement over Holt time series forecasting methodology; it considers estimated levels, trends as well as seasonality for a h-step ahead forecasting. Thus, it consists of level *l*_*t*_, trend *b*_*t*_ and seasonality *s*_*t*_ with smoothing parameters *α, β* and *γ* respectively. There are two variations of Holt-Winters seasonal methods: the additive and multiplicative.

For time *t with m* frequency of seasonality *the* Holt-Winters additive method is:

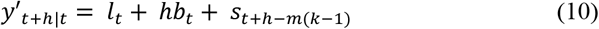

Estimated values of *level l*_*t*_, *trend b*_*t*_ and seasonality *s*_*t*_ are mathematically expressed as equations 11, 12 and 13 respectively.

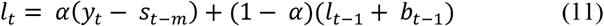

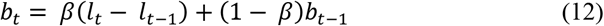

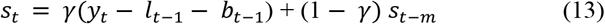

The Holt-Winter multiplicative variant can be represented as

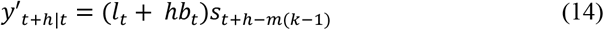

The mathematical expressions of the estimated values of *level l*_*t*_, *trend b*_*t*_ and seasonality *s*_*t*_ are expressed as equations 15, 16 and 17 respectively.

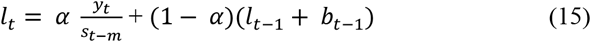

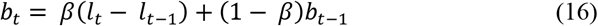

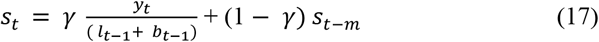

It has been shown that the additive Holt-Winter has an advantage when the seasonality component is roughly constant in the time series, while multiplicative is preferred when seasonality in the series is changing in proportional to its level. (Hyndman & Athanasopoulos, 2018).

### 3.4 Evaluation

For each of the states we forecasted the total number of people that may contract COVID-19 as well as the number who would be Blacks. Also, for each state, we forecasted the number of those who will die if they contract COVID-19, also forecast was done for the Blacks who may die of COVID-19. Holt and Holt-Winters were used as our forecasting modeling. Performance evaluation was done with Mean Absolute Percentage Error (MAPE). The forecasting error *e*_*t*_ is given as the difference between the estimated value *y*′_*t*_ and the actual value *y*_*t*_,

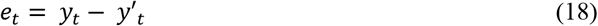

*p*_*t*_ is the percentage ratio between the error of the model and the actual value.

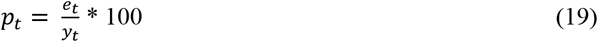

The MAPE is the absolute mean of *p*_*t*_

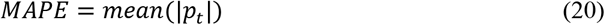

The MAPE has been effectively used in evaluating the accuracy of forecasting models. In predicting infant mortality rate, Purwanto et.al. compared the effectiveness of ARIMA, Neural Network and Linear Regression using MAPE (Purwanto, Eswaran, & Logeswaran, 2010).

Results of our forecast are shown graphically in figures 5 to 12. As shown the graphs are divided into 2020 and 2021. The forecast is for the first quarter of 2021. As proposed, we experimented with Holts and Holts-Winters exponential smoothing forecast methodologies. For each state in our study, we forecasted for total cases and death. We also forecasted for Black cases and deaths. We compared the performance of the two forecasting models in each of the state. Performance evaluation was based on MAPE. The effectiveness of COVID-19 forecast modeling using MAPE has been shown (Oladunni, Denis, Ososanya, & Barry, 2021). The performance comparison table is shown in table 2. The table shows that in most of the states, Holts-Winters exponential smoothing outperformed the Holts exponential smoothing. This suggests that seasonality is a factor.

**Figure 5.**
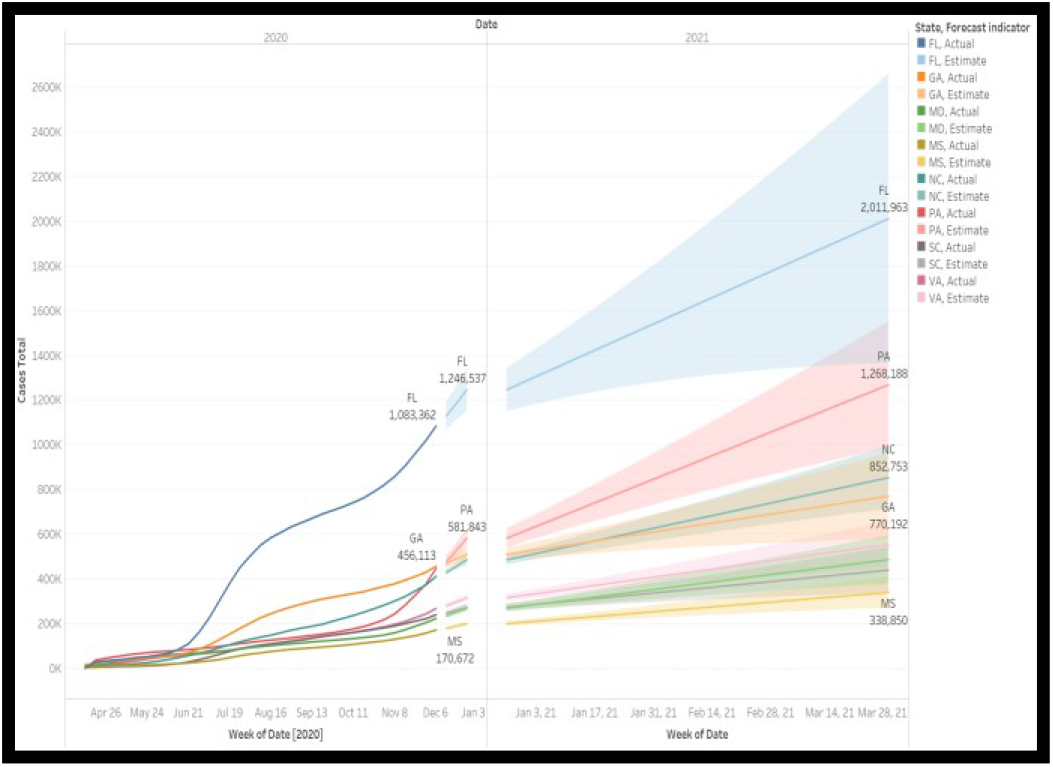
Holt exponential forecast of total COVID 19 Cases in FL, GA, MD, NC, SC, MS, PA and VA

**Figure 6.**
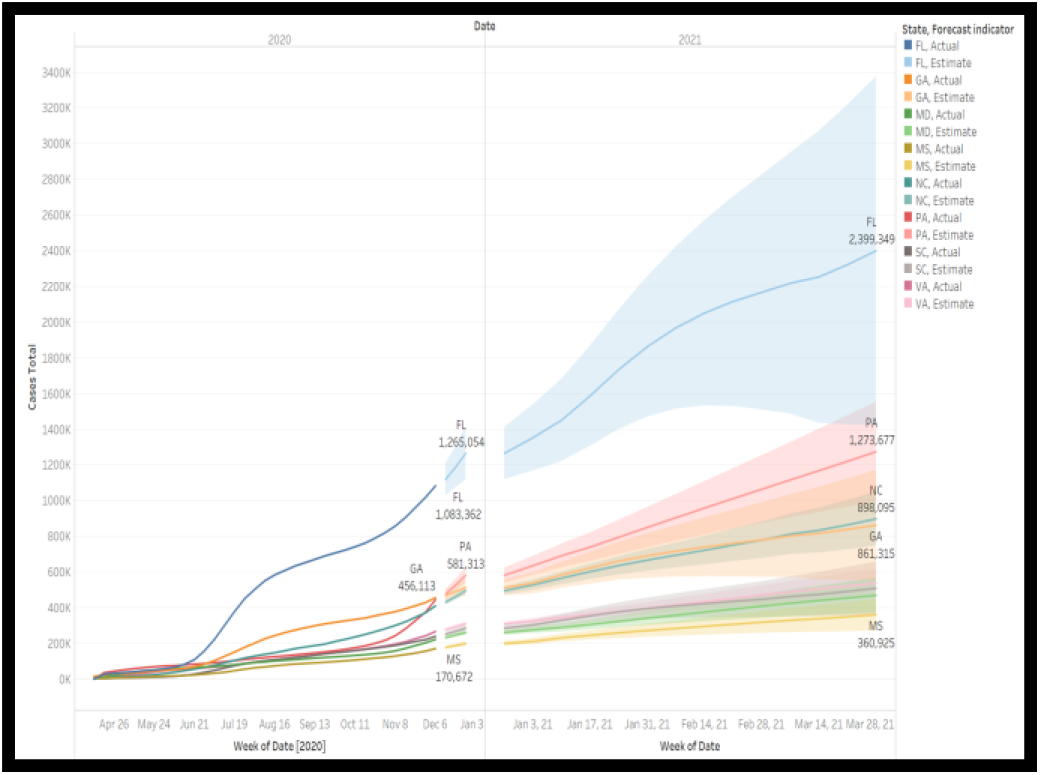
Holt-Winter exponential forecast of total COVID 19 Cases in FL, GA, MD, NC, SC, MS, PA and VA

**Figure 7.**
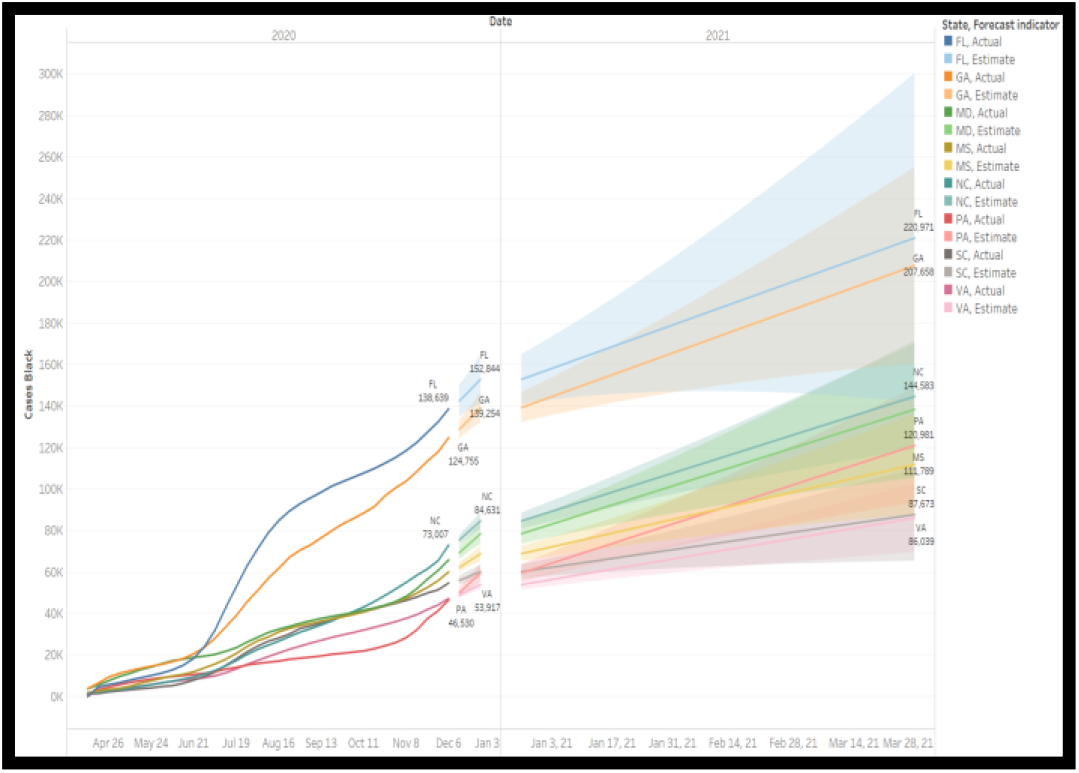
Holt exponential forecast of Black COVID 19 Cases in FL, GA, MD, NC, SC, MS, PA and VA

**Figure 8.**
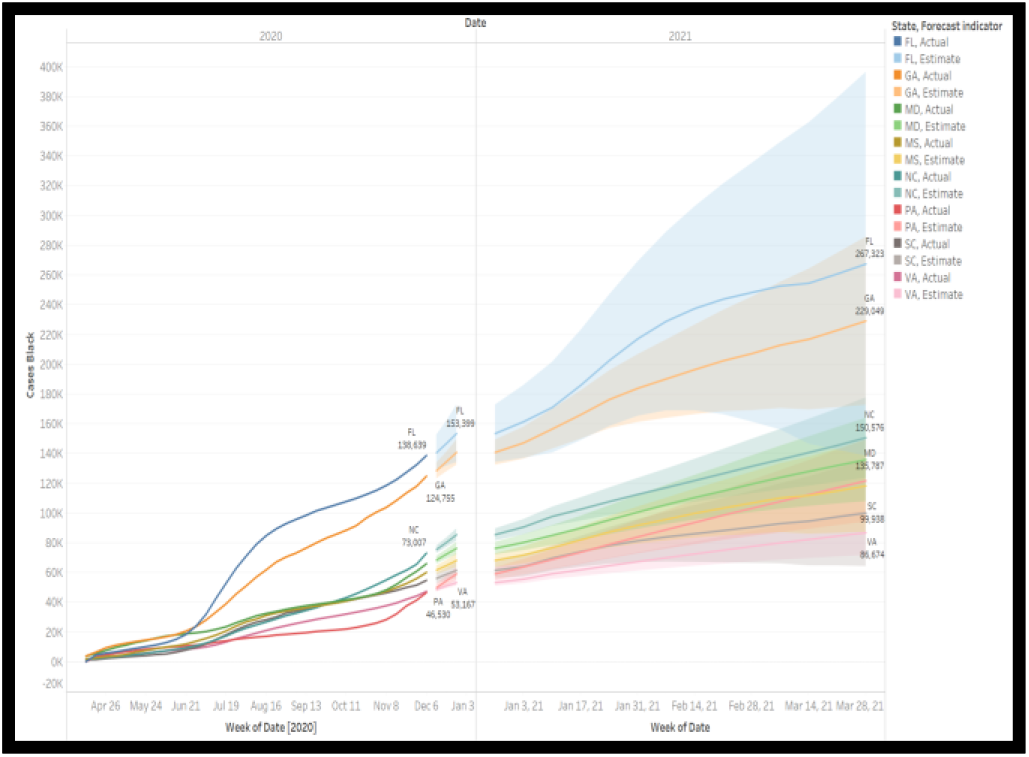
Holt-Winter exponential forecast of Black COVID 19 Cases in FL, GA, MD, NC, SC, MS, PA and VA

**Figure 9.**
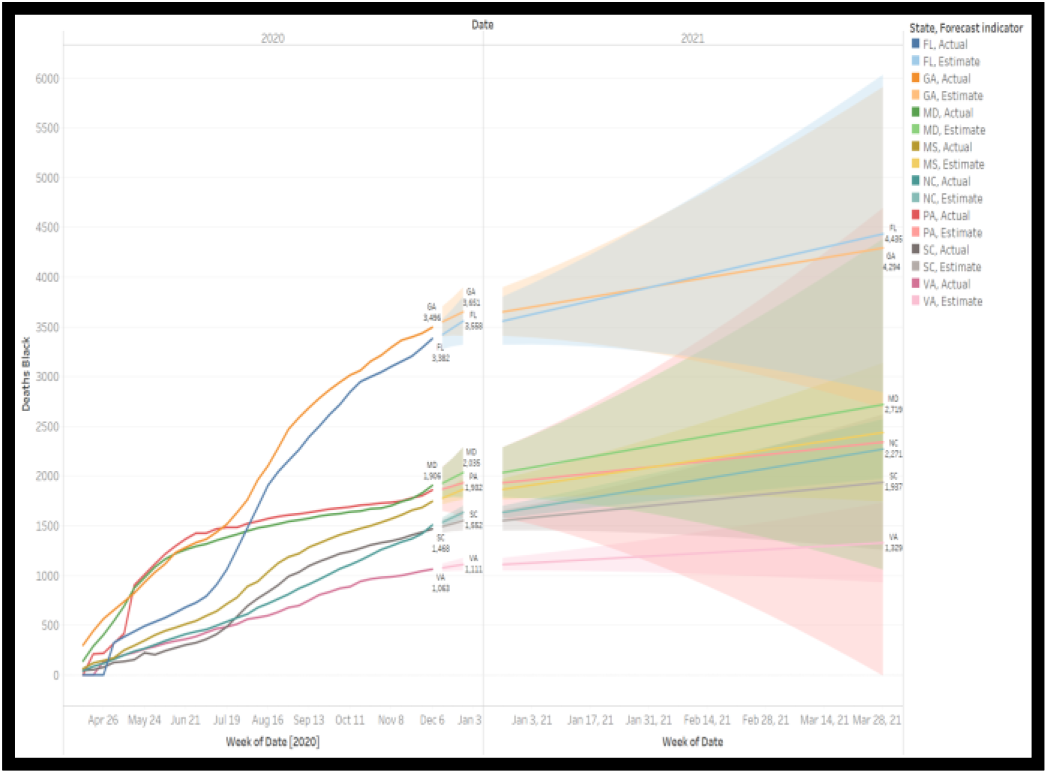
Holt exponential forecast of Black COVID 19 Deaths in FL, GA, MD, NC, SC, MS, PA and VA

**Figure 10.**
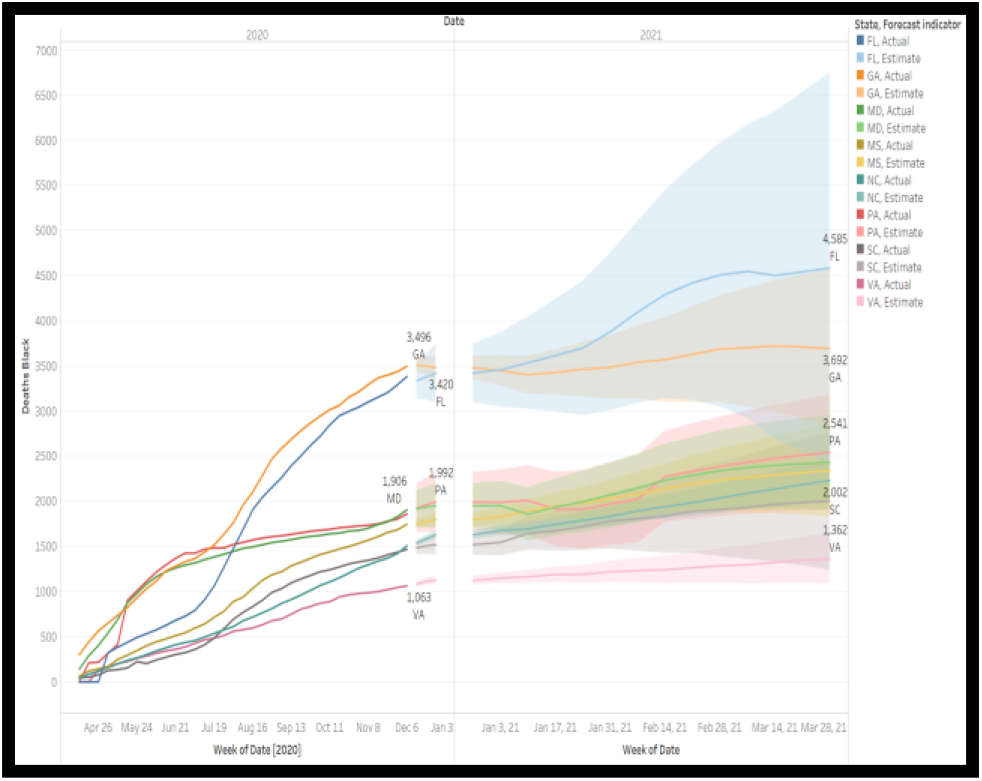
Holt-Winter exponential forecast of Black COVID 19 Deaths in FL, GA, MD, NC, SC, MS, PA and VA

**Figure 11.**
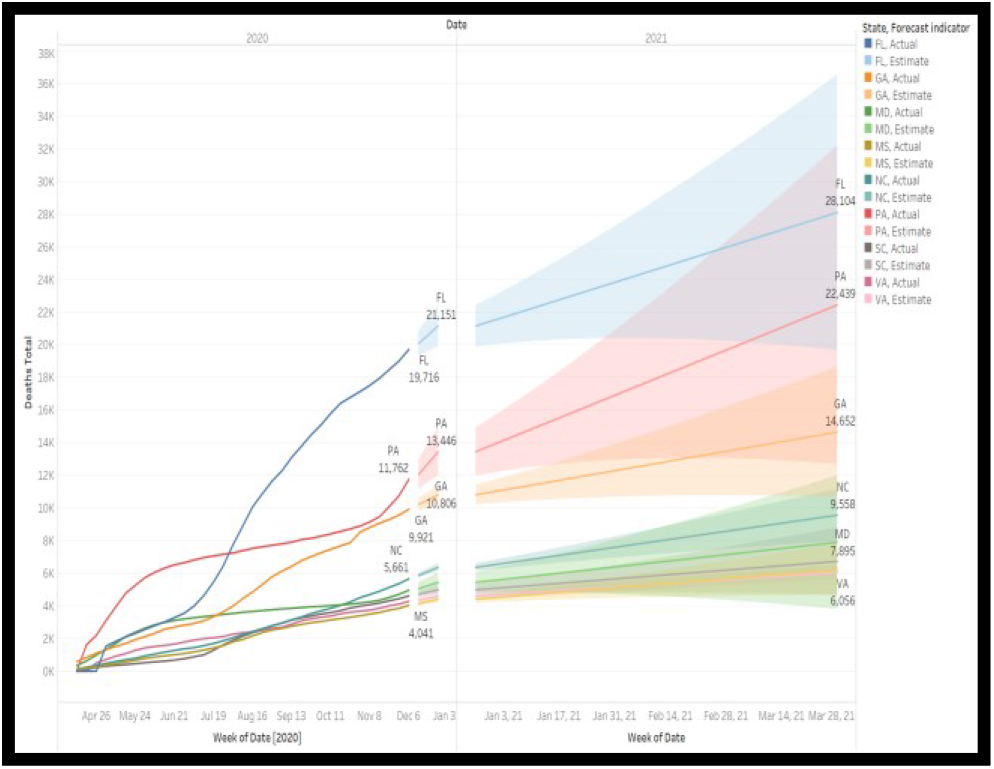
Holt exponential forecast of total COVID 19 Deaths in FL, GA, MD, NC, SC, MS, PA and VA

**Figure 12.**
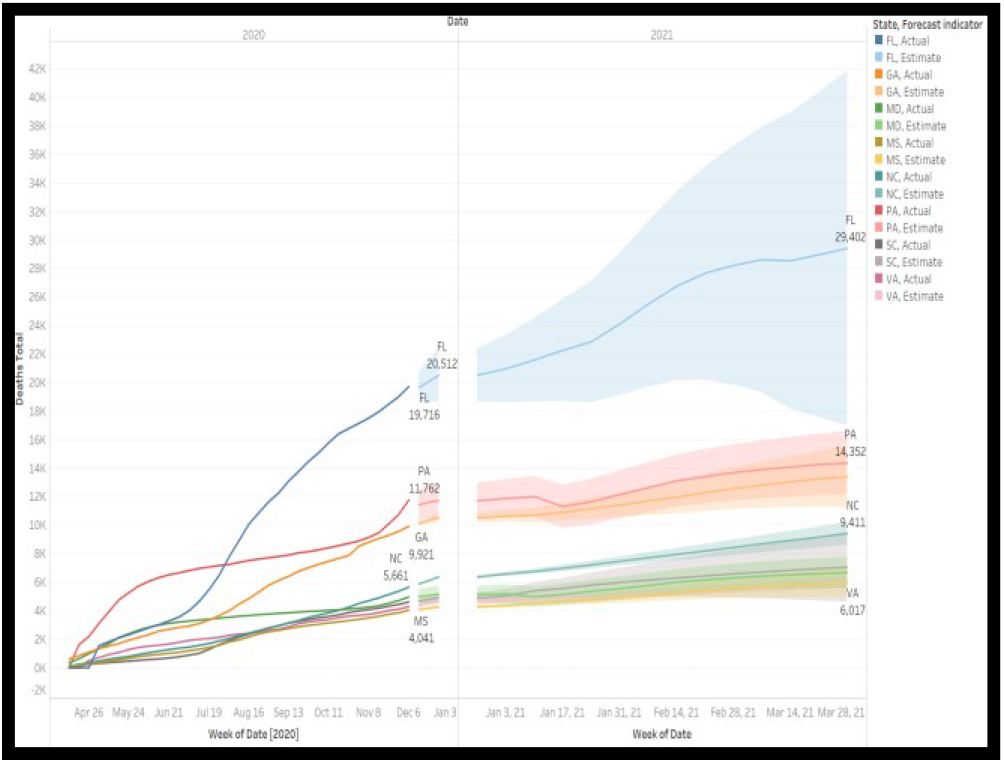
Holt-Winter exponential forecast of total COVID 19 Deaths in FL, GA, MD, NC, SC, MS, PA and VA

**Table 2.** Performance Table of Holt and Holt-Winters Forecasting Models

|  |  | VA | SC | PA | NC | MS | MD | GA | FL |
| --- | --- | --- | --- | --- | --- | --- | --- | --- | --- |
| T_C | Holt | 12.1 | 23.7 | 5.6 | 21.6 | 14.7 | 9.5 | 10.3 | 17.6 |
|  | Holt_W | 6.2 | 19.9 | 4.3 | 9.3 | 13.2 | 7.5 | 12.5 | 16.0 |
| T_D | Holt | 5.0 | 14.5 | 7.1 | 12.3 | 12.6 | 10.1 | 7.0 | 6.5 |
|  | Holt_W | 4.5 | 18.6 | 8.5 | 4.0 | 4.4 | 14.4 | 2.5 | 6.2 |
| B_C | Holt | 12.2 | 15.2 | 11.7 | 14.2 | 11.1 | 8.7 | 11.5 | 17.1 |
|  | Holt_W | 6.3 | 16.0 | 5.5 | 9.1 | 7.3 | 4.2 | 8.1 | 14.2 |
| B_D | Holt | 4.7 | 12.2 | 8.6 | 10.9 | 9.0 | 10.5 | 6.2 | 5.8 |
|  | Holt_W | 3.7 | 12.5 | 15.1 | 5.7 | 4.6 | 14.2 | 2.7 | 5.0 |

T\_C, T\_D, B\_C and B\_D are the total cases, total deaths, Black cases and Black deaths respectively.

As shown in table 2, in most of the states, except for a few outliers, the Holt-Winter (Holt_W) models have lower MAPE than the Holt models. Notably in MD, the fact that Holt outperformed Holt-Winters in forecasting deaths suggests that seasonality might not factor into the forecast of deaths in this state. The MAPE of SC on both models are in double digits; suggesting that our models did not completely capture all the underlying time series patterns of the state. A better model is defined as the one with the lower MAPE. Therefore, model selection was based on the performance on the MAPE. Using this approach, we obtained table 3.

**Table 3.** Model Selection Table

|  | VA | SC | PA | NC | MS | MD | GA | FL |
| --- | --- | --- | --- | --- | --- | --- | --- | --- |
| T_C | HW | HW | HW | HW | HW | HW | Holt | HW |
| T_D | HW | Holt | Holt | HW | HW | Holt | HW | HW |
| B_C | HW | Holt | HW | HW | HW | HW | HW | HW |
| B_D | HW | Holt | Holt | HW | HW | Holt | HW | HW |

HW and Holt are the Holt-Winter and Holt exponential forecasting models, respectively.

As shown in table 3, HW outperformed Holt in most of the cases and deaths. Table 4 shows the forecasted values from the selected models. For example, Holt Winter was selected as the better model for the total cases in FL because it has a lower MAPE as compared with Holt. The forecasted value at FL turned out to be 2, 399, 349. In SC, Holts Winter was also selected as the better model to forecast the total cases with a forecasted value of 509, 004.

**Table 4.** Forecasted Values

| State | Total Cases | Black Cases | Black Cases/Tot Cases % | Total Death | Black Death | (Black Death/Total Death) % | CHCD |
| --- | --- | --- | --- | --- | --- | --- | --- |
| FL | 2399349 | 267323 | 11.14% | 29402 | 4435 | 15.08% | 3.94% |
| GA | 770192 | 229049 | 29.74% | 13393 | 4294 | 32.06% | 2.32% |
| MD | 468683 | 135787 | 28.97% | 7895 | 2719 | 34.44% | 5.47% |
| MS | 360925 | 118198 | 32.75% | 5951 | 2438 | 40.97% | 8.22% |
| NC | 898095 | 150576 | 16.77% | 9411 | 2271 | 24.13% | 7.37% |
| PA | 1273677 | 121590 | 9.55% | 22439 | 2341 | 10.43% | 0.89% |
| SC | 509004 | 170730 | 33.54% | 6728 | 1937 | 28.79% | -4.75% |
| VA | 537892 | 86674 | 16.11% | 6017 | 1329 | 22.09% | 5.97% |

As shown in table in table 4, *we do not find enough evidence against our hypothesis. Therefore. we contend that CHCD would have continue in the black community*.

## 4. PREDICTING COVID-19 FATALITY RATE IN AFRICAN AMERICAN COMMUNITY

Using time series analysis and forecasting modeling, we have shown that there is a strong evidence of disproportionality in the impact of the coronavirus pandemic in the African American community. This disproportionality in the impact suggests that a universal modeling of the pandemic in the US may be inadequate to predict fatality rate in the Black community. Therefore, the next stage of our investigation is to design, develop and evaluate a COVID-19 fatality predictive model with the capability of predicting fatalities in the Black community.

Dataset for this experiment was obtained from the John Hopkins COVID-19 repository (John Hopkins University, n.d.), US Census Bureau (US Census Bureau, n.d.) and US Center for Disease Control and Prevention (CDC) (Center for Disease Control and Prevention, n.d.). The data consisted of the fatality rates of the pandemic in each county of the US. Since the focus our work is on the Black community, we targeted counties that we considered as a true representation of the Black community. We assumed a Black county to be any county in the US that are at least 45% Black. Based on the number of ethnic groups in the US, we believe that this is a fair assumption. Figure 13 shows the geographical location of these counties on the US map. The experimental design flowchart is shown in figure 14.

**Fig 13.**
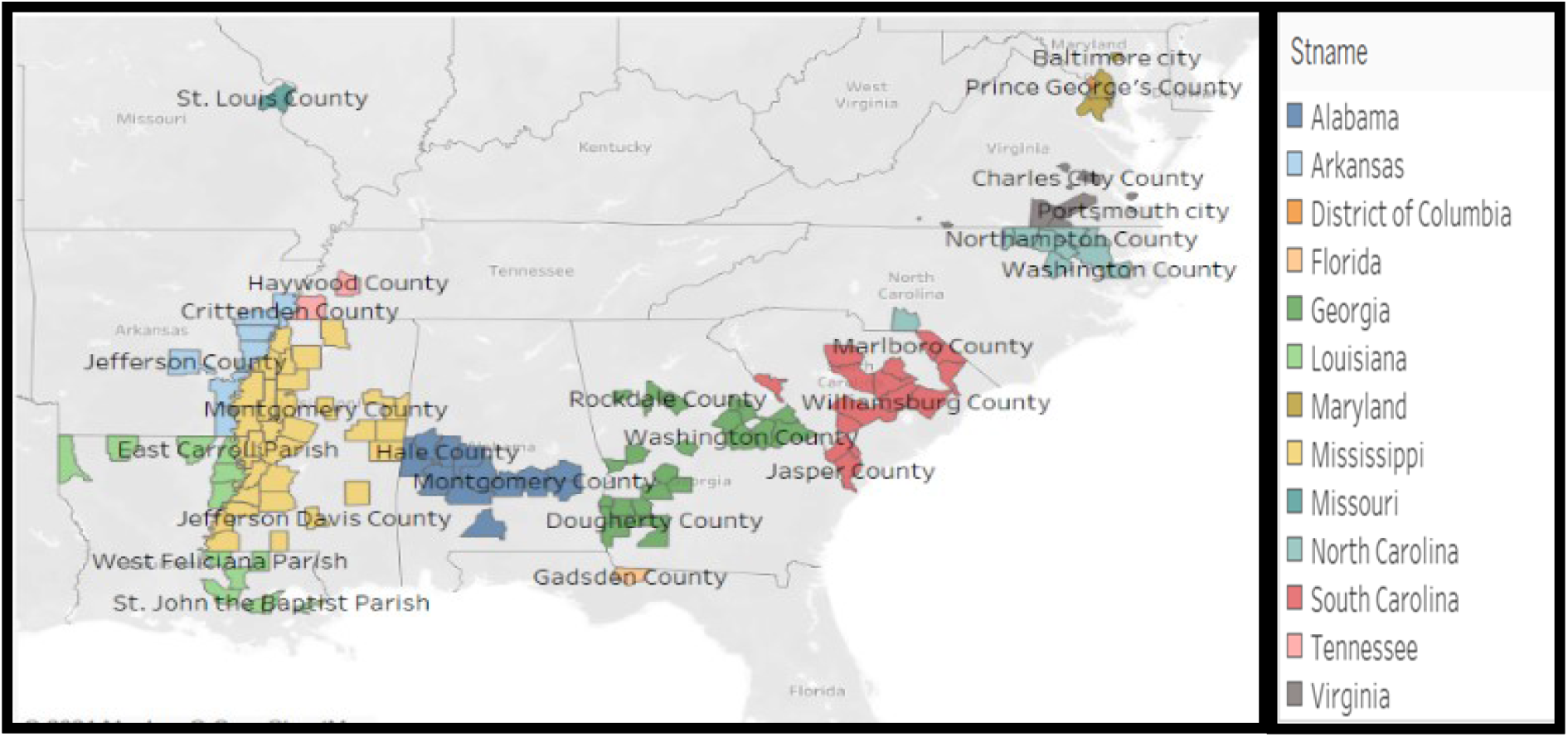
A modeled geographical map of counties with at least 45% Black population in the USA

**Figure 14.**
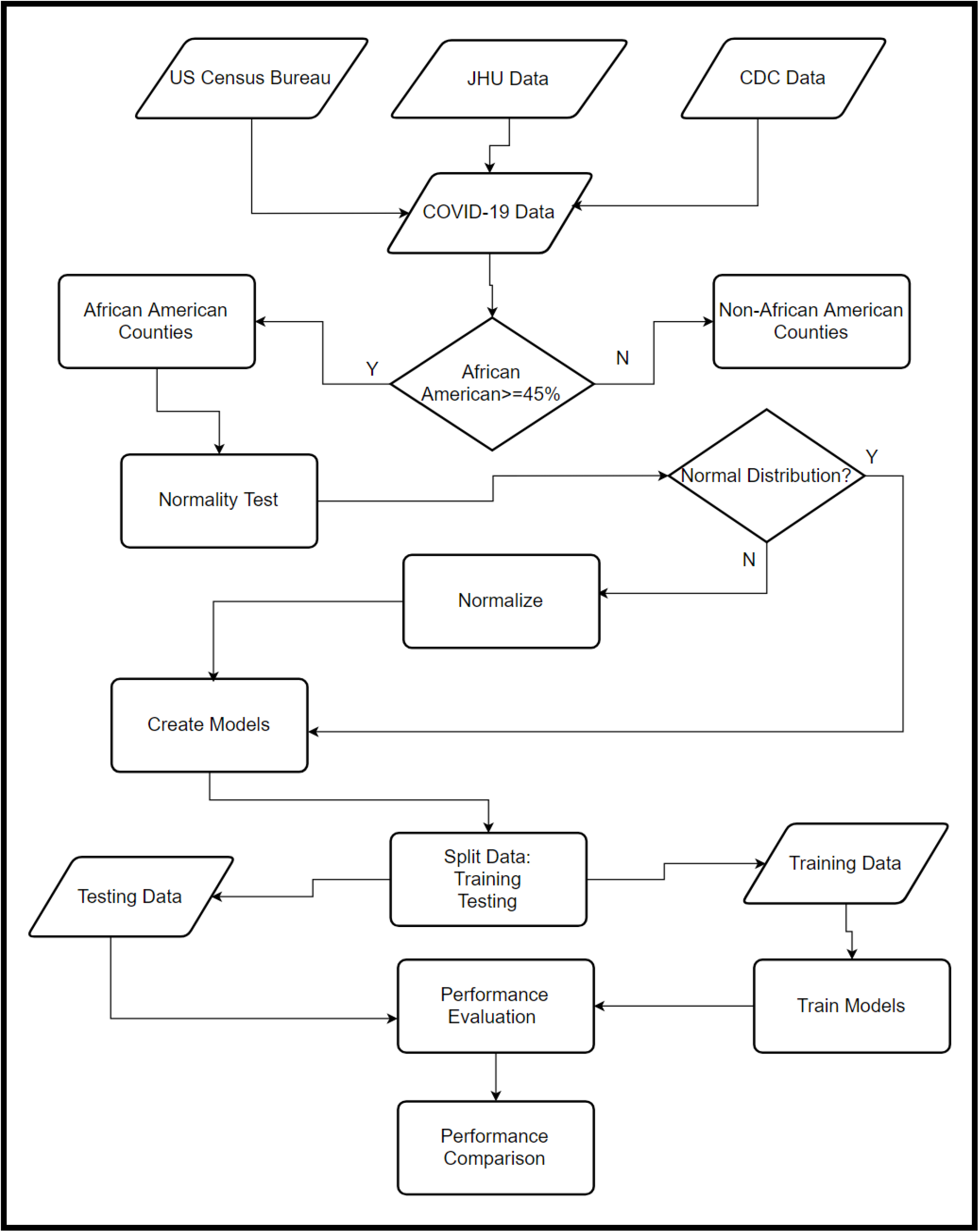
experimental Design Flowchart

### 4.1 Z Standardization

Dataset consisted of predictors with varying degree of magnitudes. Therefore, feature scaling was done to make all predictors be on the same scale. This ensures that all predictors are equally important. Min-max normalization is a very popular normalization approach; however, Z standardization is preferred because of its ability to handle outliers.

Mathematically,

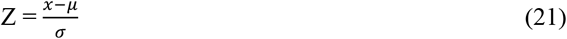

Where z, *x, μ* and *σ* are the normalized values, predictor, mean and standard deviation, respectively.

Dataset was then split into 75% and 25% for training and testing, respectively.

### 4.2 Learning Algorithm

Five different learning algorithms were trained and evaluated to predict fatality rate in the Black community, namely: Linear Regression, Decision Tree, K-Nearest Neighbor (KNN), and Support Vector Machine (SVM) model. We evaluated the performances of the models using Mean Square Error (MSE).

#### 4.2.1 Linear Regression

Linear Regression maps a linear relationship between a response variable *Y* and feature vectors **X**.

A simple linear regression can be represented as

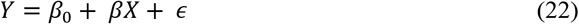

To accommodate *p* number of predictors X, a simple linear regression is modelled to multiple linear regression.

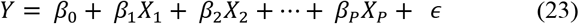

The regression coefficients are estimated as

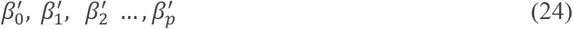

Then

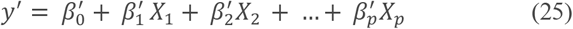

Where *y*′ is the estimated response variable.

Therefore, the Residual Sum of Squares error is

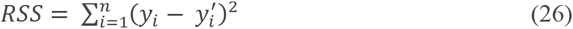

Substituting equation 25 into equation 26

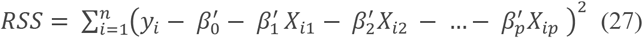

For this study, ridge regression was used. The ridge regression is a variant of the multiple linear regression with a shrinkage penalty parameter λ. Additional constrain was added to RSS

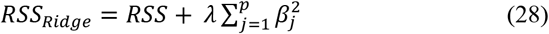

As shown in equation 28, the ridge regression seeks to improve the fitness of the multiple linear regression with the additional penalty λ. The penalty term has no effect when λ =0, which produces the multiple linear regression. However, the impact of the penalty grows as λ -> ∞ and the ridge coefficient estimates tends to zero. For this model, the value of λ was set to 1.0E-8. Feature selection strategy was used to determine all necessary and sufficient predictors for the ridge regression. Our feature selection strategy was based on the M5 Prime. The M5 prime feature selection model is a variant of decision tree that builds multiple linear regressions at each node. Predictors with a low standardized value are eliminated to improve the Akaike Information Criterion (AIC).

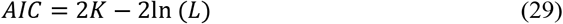

Where k and L are the predictors and likelihood of the models respectively.

#### 4.2.2 Decision Tree

A Decision Tree is a tree where every node addresses a test of a predictor and the leaf node gives classification or regression. In this study, we built a regression decision tree. The test model is characterized by beginning at the root, testing attribute values at every node, and arranging down to the suitable branch till it arrives at the leaf node which gives classification.

Suppose *X*_*1*_, *X*_*2*_, *…, X*_*P*_ are *P* predictors in a feature vector *X*. The goal of a decision tree is to divide them into a predictor space *j* distinct and *R*_*1*_, *R*_*2*_, *…, R*_*J*_ non-overlapping regions. A decision tree model is optimized when the Residual Sum of Squares error *RSSE* is at the minimum. Given a cutpoint *s*, a recursive binary splits the predictor space into {*X*|*X*_*j*_| < *s*} and {*X*|*X*_*j*_| ≥ *s*} to obtain the minimum *RSSE*.

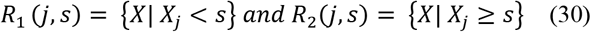

An optimized decision tree seeks j and s that minimized.

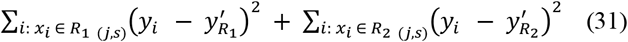

Where 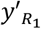 and 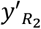 are the mean estimated response for the regions *R*_*1*_ and *R*_*2*_ respectively.

In this study, 0.01, 2 and 4 were selected as the hyperparameter values for minimal gain, minimal leaf size and minimum size for split respectively.

#### 4.2.3 Support Vector Machine (SVM)

The SVM sets up an isolating hyperplane and a maximal margin by picking a subset SV⊂X called support vectors. The optimization problem is given by Equation 32. The SVs are utilized to compute the normal vector W on the hyperplane and the bias b to satisfy the requirement on the optimization problem.

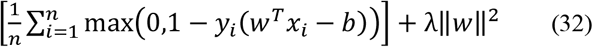

where ***y***_***i***_ is the class to which point ***x***_***i***_ belongs, **w** is the normal vector, and λ is a tuning parameter. During the implementation of this model, we used the dot kernel type, a kernel cache of 200, a convergence epsilon of 0.001, a max iteration of 100000 and a L pos and L neg of 1.0 (L pos and L neg are SVM complexity constants).

#### 4.2.4 K-Nearest Neighboring (KNN)

KNN is known to be basic and simple learning algorithm. This algorithm is executed by looking for the group of K folds, in the nearest training data (similar) objects in new data or testing data. For the most part, the Euclidean distance formula is utilized to characterize the distance between datapoints (Okfalisa, 2017). The Euclidean distance is mathematically represented as

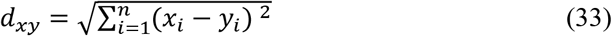

Where d represents the distance, x and y are 2 data points.

For this experiment 10 was chosen as the value of K, in addition we used the MixedMeasures for the measure types.

#### 4.2.5 Neural Network (NN)

A Neural Network (NN) is a set of interconnected artificial neurons networked after the human brain. It has the capability of pattern recognition and knowledge discovery in a dataset. In a simplest form, it consists of a neuron (perceptron). A more complex network comprises of several layers of neurons. The multilayer feedforward network or multilayer perceptron is partitioned into three layers: input layer, hidden layer, and output layer. For this study, our NN comprises of 2 hidden layers with a training cycles of 200, a learning rate of 0.01, a momentum of 0.9, and an error epsilon of 1.0E-4.

### 4.3 Performance *Comparison*

For a better analysis and model comparison of the results of our experiment, we will evaluate their performances using the following criteria.

#### 4.3.1 Root Mean Squared Error (RMSE)

RMSE shows the prediction errors, defining the spread of the residual, and how concentrated the data is around the line of best fit.

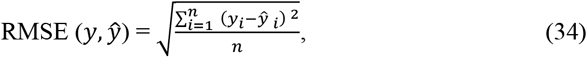

Where *n is the number of examples, y*_*i*_, *and ŷ*_*i*_ are true and predicted values of the response variables respectively.

#### 4.3.2 Relative Error (RE)

When used as a measure of precision, it is the ratio of the absolute error of the actual value to the predicted value.

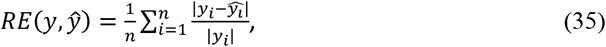

where |***y***_***i***_ − ***ŷ***_***i***_ | is the absolute error. |***y***_***i***_| and |***ŷ***_***i***_| are the absolute values of the actual and predicted respectively. n is the number of records.

#### 4.3.3 Absolute error (AE)

It is the absolute difference between the predicted value of a quantity ***x***_***0***_ and its actual value x.

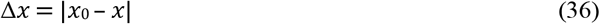

#### 4.3.4 Relative Error Lenient (REL)

The average of the absolute deviation of the predicted value from the actual value divided by the maximum of the absolutes actual value and predicted value.

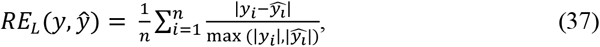

where |***y***_***i***_ − ***ŷ***_***i***_ | is the absolute error, |***y***_***i***_| is the actual value, |***ŷ***_***i***_| is the predicted value and n the number of errors.

#### 4.3.5 Relative Error Strict (RES)

RES is the average of the absolute deviation of the prediction from the actual value divided by the minimum of the actual value and the predicted value

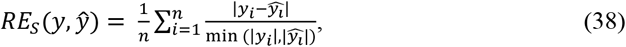

Where |***y***_***i***_ − ***ŷ***_***i***_ | is the absolute error, |***y***_***i***_| is the absolute actual value, |***ŷ***_***i***_| is the absolute predicted value and n the number of errors.

#### 4.3.6 Root Relative Squared Error (RRSE)

RRSE criterion measures the relative error of what the error would have been if a simple predictor had been used. Thus, normalizing the total squared error.

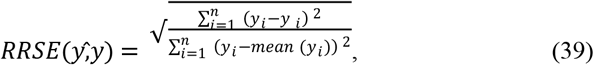

where ***y***_***i***_ the taken measure and ***ŷ***_***i***_ the prediction value.

#### 4.3.7 Squared Error (SE)

It is the average squared difference between the estimated values and the actual value.

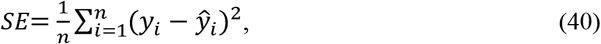

where ***y***_***i***_ and ***ŷ***_***i***_ are the actual and predicted values respectively.

#### 4.3.8 Correlation

Correlation shows a linear relationship between two variables.

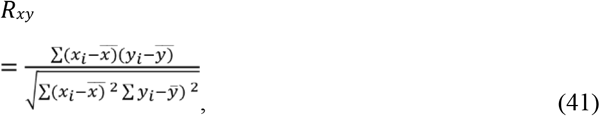

where *R*_*xy*_ is correlation coefficient, ***x***_***i***_ is the *i*^*th*^ values of the x variable in a sample, 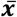 is mean of the values of the x variable, *y*_*i*_ is the *i*^*th*^ value of the y-variable in a sample, and 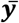 is the mean of the values of the y variable.

#### 4.3.9 Squared Correlation

Squared correlation represents the proportion of the variance for a dependent variable that is explained by an independent variable or variables in a regression model

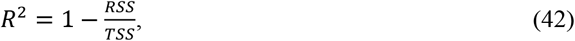

where *R*^2^ is the Squared correlation, RSS= Sum of squares of residuals, TSS= Total sum of squares

#### 4.3.10 Prediction average

This is the mean value of predictions. The sum of all predicted values is divided by the number of all predictions. Using the above criteria, table 5 shows the performance comparison of the models.

**Table 5.** Models’ Performance Comparison

| Criteria | Ridge Regression | Decision Tree | SVM | K-NN | NN |
| --- | --- | --- | --- | --- | --- |
| Root mean squared error. | 4.279 | 0.825 | 1.462 | 1.659 | 1.125 |
| Absolute error | 2.656 | 0.627 | 1.121 | 1.308 | 0.935 |
| Relative error strict | 121.00 | 22.29 | 48.12 | 53.74 | 42.77 |
| Relative error lenient | 42.15 | 17.77 | 28.22 | 31.36 | 26.03 |
| Root relative squared error | 2.933 | 0.520 | 0.894 | 1.022 | 0.740 |
| Squared error | 50.110 | 0.754 | 2.279 | 2.976 | 1.325 |
| Correlation | 0.220 | 0.881 | 0.655 | 0.311 | 0.863 |
| Squared correlation | 0.132 | 0.781 | 0.461 | 0.169 | 0.751 |
| Prediction average | 3.288 | 3.288 | 3.288 | 3.288 | 3.288 |

As it is shown in Table 5, based on all evaluation criteria, the Decision Tree model, which has the lowest RMSE = 0.83, is the model that fits best for this study. The implementation of these models was done in Python, Tableau and RapidMiner.

## 5. CONCLUSION

In this study we have analyzed the impact of COVID-19 in the African American community. Time series analysis were used to show the disproportionality in COVID-19 impact. Visualization of the trajectory of the coronavirus pandemic was shown using area graphs. For a better understanding of the time series, timelines were shown in months as well as in quarters of 2020. We studied the trajectory of total cases, Black cases, total deaths, and Black deaths. The time frame of our work spans March 13 to December 16, 2020. We computed the COVID-19 Health Care Disproportionality for the time frame.

Furthermore, we designed, developed and evaluated eight COVID-19 forecasting models using Holts and Holts-Winters exponential smoothing forecasting methodologies. Forecast was made for the total cases, Black cases, total deaths and Black deaths. Using MAPE, we built a model selection table containing the best forecasting results. A forecast table was then built for total cases, Black cases, total deaths and Black deaths showing COVID-19 health care disproportionality for each state. The results of our forecast modeling suggests that COVID-19 Health Care Disproportionality will continue to the end of the first quarter of 2021.

As shown in the study, our experimental result suggests that there exists a strong evidence of COVID-19 disproportional impact in the Black community. We argued that a universal modeling of COVID-19 fatality rate in the US is inadequate to predict fatality rate in the Black community. Therefore, a fatality rate predictive model was designed, developed and evaluated specifically for the Black counties. Based on the US ethnic composition, we assumed a Black county to be a county with at least 45% Black population. Decision Tree, Support Vector Regression, Neural Networks, K-Nearest Neighbors and Ridge Regression learning algorithms were trained and evaluated. The outcome of our experiment showed that the Decision Tree model had the best performance in predicting fatality rates in a Black county.

## Data Availability

Dataset is available at COVID Racial Data Tracking Project, CDC, US Census Bureau and John Hopkins COVID 19 Repository

https://covidtracking.com/race

https://covid.cdc.gov/covid-data-tracker/#datatracker-home

https://coronavirus.jhu.edu/

https://www.census.gov/data.html

## 6. IMPLICATION OF STUDY

This study has the following implications:

1. The coronavirus pandemic disproportionally impacted the Black community.
2. Decision Tree has the best performance in modelling fatality rate in the Black Community. The tree suggested that Black and senior citizens with pre-existing condition living in Georgia State are the most vulnerable.
3. If healthcare disproportionality continues, the impact of the next pandemic in the Black community should be a concern to all stakeholders.

## 7. LIMITATION OF STUDY

Since study was limited to selected states and counties, experimental results may be different in states that do not have a large population of African Americans.

## 8. FUTURE WORK

Our future work includes the following:

1. Parameterize the effect of COVID-19 vaccination in the Black community; showing how much the vaccine affected the prognosis of the pandemic.
2. Develop a framework for a universal pandemic preparedness.

## 9. AKNOWLEDGMENT

This work is funded by the National Science Foundation grant number 2032345.

## References

Ahmad, S., & Latif, H. A. (2011). Forecasting on the crude palm oil and kernel palm production: Seasonal ARIMA approach. IEEE Colloquium on Humanities, Science and Engineering. Penang.

Andersen, M. (2020). Early Evidence on Social Distancing in Response to COVID-19 in the United States. Greenboro: UNC.

Anderson, M. (2016, April 07). Who relies on public transit in the U.S. Retrieved January 05, 2021, from https://www.pewresearch.org/fact-tank/2016/04/07/who-relies-on-public-transit-in-the-u-s/

Atlantic, T. (2020). The COVID Tracking Project. Retrieved Novemeber 22, 2020, from https://covidtracking.com/race

Branigin, A. (2020, March 31). Black Communities Are on the ‘Frontline’ of the COVID-19 Pandemic. Here’s Why. Retrieved January 05, 2020, from https://www.theroot.com/black-communities-are-on-the-frontline-of-the-covid-19-1842404824

Center for Disease Control and Prevention. (n.d.). COVID Data Tracker. Retrieved from https://covid.cdc.gov/covid-data-tracker/#datatracker-home

Centers for Disease Control and Prevention. (n.d.). Evidence for Limited Early Spread of COVID-19 Within the United States, January–February 2020. Retrieved December 10, 2020, from https://www.cdc.gov/mmwr/volumes/69/wr/mm6922e1.htm

Centers for Disease Control and Prevention. (n.d.). Asthma Surveillance Data. Retrieved December 10, 2020, from https://www.cdc.gov/asthma/asthmadata.htm

Cneters for Disease Control and Prevention. (n.d.). HIV and African Americans. Retrieved December 10, 2020, from https://www.cdc.gov/hiv/group/racialethnic/africanamericans/index.html

Cunningham, T. J., Croft, J. B., Liu, Y., Lu, H., Eke, P. I., & Giles, W. H. (2017). Vital signs: racial disparities in age-specific mortality among blacks or African Americans—United States, 1999–2015. MMWR Morb Mortal Wkly Rep.

Dowd, J. B., Andriano, L., Brazel, D. M., Rotondi, V., Block, P., Ding, X., … Mills, M. C. (2020). Demographic science aids in understanding the spread and fatality rates of COVID-19. National Academy of Sciences.

Dutko, P., Ploeg, M. V., & Farrigan, T. (August 2012). Characteristics and influential factors of food deserts. Washington, DC:: US Department of Agriculture, Economic Research Service,.

Dyer, O. (2020). “Covid-19: Black people and other minorities are hardest hit in US,. BMJ, 369.

Elmunim, N. A. M. Abdullah, A. M., & Zaharim, A. (2015). Forecasting ionospheric delay during quiet and disturbed days using the Holt-Winter method. International Conference on Space Science and Communication (IconSpace). Langkawi.

Ferdinand, K. C., & Samar A. Nasser. (2020). African-American COVID-19 Mortality: A Sentinel Event. Journal of the American College of Cardiology, 2746–2748.

Fletcher, F. E., Allen, S., Vickers, S. M., Beavers, T., Hamlin, C. M., Young-Foster, D., … Erwin, P. C. (2020). COVID-19’s Impact on the African American Community: A Stakeholder Engagement Approach to Increase Public Awareness Through Virtual Town Halls. Health Equity, 4(1).

Hyndman, R. J., & Athanasopoulos, G. (2018). Forecasting: Principles and Practice. Melbourne, Australia: OTexts. Retrieved from https://otexts.com/fpp2/translations.html

John Hopkins University. (n.d.). Retrieved from https://coronavirus.jhu.edu/about/how-to-use-our-data

John Hopkins University. (n.d.). Coronavirus Resource Center. Retrieved January 11, 2021, from https://coronavirus.jhu.edu/map.html

Kochar, R., & Cilluffo, A. (2017, Novemeber 01). Pew Review Center. Retrieved January 05, 2020, from https://www.pewresearch.org/fact-tank/2017/11/01/how-wealth-inequality-has-changed-in-the-u-s-since-the-great-recession-by-race-ethnicity-and-income/

Laurencin, C. T., & McClinton, A. (2020). The COVID-19 Pandemic: a Call to Action to Identify and Address. Journal of Racial and Ethnic Health Disparities (, 7, 398–402.

M., H. (1985). Report of the Secretary’s Task Force on Black & Minority Health.. Washington, DC: U.S. Department of Health and Human Services,.

Mege, C. R., Haq, I. N., Leksono, E., & Soelami, F. X. (2019). Battery Discharging Temperature Prediction Using Holt’s Double Exponential Smoothing. 6th International Conference on Electric Vehicular Technology (ICEVT). Bali, Indonesia.

Morens, D. M., Breman, J. G., Calisher, C. H., Doherty, P. C., Hahn, B. H. T. Keusch, G., … Taubenberger, J. K. (2020). The Origin of COVID-19 and Why It Matters. The American Journal of Tropical Medicine and Hygiene, 103(3), 955.

nature. (n.d.). Meet the scientists investigating the origins of the COVID pandemic. Retrieved December 10, 2020, from https://www.nature.com/articles/d41586-020-03402-1

Navarro, M. M., & Navarro, B. B. (2019). Optimal Short-Term Forecasting Using GA-Based Holt-Winters Method. IEEE International Conference on Industrial Engineering and Engineering Management (IEEM). Macao.

New York Times. (2021, January 07). Maryland Coronavirus Map and Case Count. Retrieved January 07, 2021, from https://www.nytimes.com/interactive/2020/us/maryland-coronavirus-cases.html

Okfalisa, I. G. (2017). Comparative analysis of k-nearest neighbor and modified knearest neighbor algorithm for data classification. 2nd International conferences on Information Technology, Information Systems and Electrical Engineering (ICITISEE). Yogyakarta, Indonesia,.

Oladunni, T., Denis, M., Ososanya, E., & Barry, A. (2021). Exponential Smoothening Forecast of African Americans’ COVID-19 Fatalities. IEEE International Conference on Computing and Data Science, (pp. 466–471). Stanford, CA.

Purwanto, D., Eswaran, C., & Logeswaran, R. (2010). A Comparison of ARIMA, Neural Network and Linear Regression Models for the Prediction of Infant Mortality Rate. Fourth Asia International Conference on Mathematical/Analytical Modelling and Computer Simulation. Bornea.

R. Anggrainingsih, Prabanuadhi, A., & Yohanes, S. P. (2018). International Conference on ICT for Rural Development (IC-ICTRuDev. Forecasting The Number of Patients at RSUD Sukoharjo Using Double Exponential Smoothing Holt. Indonesia.

R.B. Hawkins, E.J. Charles, & J.H. Mehaffeyab. (2020). Socio-economic status and COVID-19–related cases and fatalities. Public Health, 189, 129–134.

Rab, S., Javaid, M., & Haleem, A. (2020). Face masks are new normal after COVID-19 pandemic. ScienceDirect, 14(6), 1617–1619.

Richardson, S., Hirsch, J. S., & Narasimhan, M. (2020). Presenting characteristics, comorbidities, and outcomes among 5700 patients hospitalized with COVID-19 in the New York city area. JAMA.

Shigute, Z., Mebratie, A. D., Alemu, G., & Bedi, a. A. (2020). Containing the spread of COVID-19 in Ethiopia. Journal of Global Health.

Ssentongo, P., Ssentongo, A. E., Heilbrunn, E. S., Ba, D. M., & Chinchilli, V. M. (2020). Association of cardiovascular disease and 10 other pre-existing comorbidities with COVID-19 mortality: A systematic review and meta-analysis. PLoS ONE, 15(8).

State Health Facts. (2019). Poverty Rate by Race/Ethnicity. Retrieved January 05, 2020, from https://www.kff.org/statedata/

Tai, D. B., Shah, A., Doubeni, C. A., Sia, I. G., & Mark L Wieland. (2020). The Disproportionate Impact of COVID-19 on Racial and Ethnic Minorities in the United States,. Clinical Infectious Diseases.

Trivedy, C., Mills, I., & Dhanoya, O. (2020). The impact of the risk of COVID-19 on Black, Asian and Minority Ethnic (BAME) members of the UK dental profession. British Dental Journal, 228, pages 919–922.

U.S. Department of Health and Human Services Office of Minority Health. Diabetes and African Americans. (n.d.). Diabetes and African Americans. Retrieved Decemeber 10, 2020, from https://minorityhealth.hhs.gov/omh/browse.aspx?lvl=4&lvlid=18

U.S. Department of Health and Human Services Office of Minority Health. Heart Disease and African Americans. (n.d.). Heart Disease and African Americans. Retrieved December 10, 2020, from https://minorityhealth.hhs.gov/omh/browse.aspx?lvl=4&lvlid=19

U.S. Department of Health and Human Services Office of Minority Health. Obesity and African Americans. (n.d.). Obesity and African Americans. Retrieved December 10, 2020, from https://minorityhealth.hhs.gov/omh/browse.aspx?lvl=4&lvlid=25

US Census Bureau. (n.d.). Retrieved from https://www.census.gov/data/datasets.html

Wells, C. R., Sah, P., Moghadas, S. M., Pandey, A., Shoukat, A., Wang, Y., … Galvani, A. P. (2020). Impact of international travel and border control measures on the global spread of the novel 2019 coronavirus outbreak. National Academy of Sciences.

WHO Director-General’s opening remarks at the media briefing on COVID-19 - 11 March 2020. (n.d.). Retrieved December 10, 2020, from https://www.who.int/director-general/speeches/detail/who-director-general-s-opening-remarks-at-the-media-briefing-on-covid-1911-march-2020

Xia, L., Yaomei, G., & Weiwei, S. (2010). Forecast to Textile and Garment Exports Based on Holt Model. International Conference of Information Science and Management Engineering. Xi’an.

Y. Guo X. S., & Zhang, X. (2010). A study of short term forecasting of the railway freight volume in China using ARIMA and Holt-Winters models,. 8th International Conference on Supply Chain Management and Information,. Hong Kong.

Yan-ming Yang, Yu, H., & Sun, Z. (2017). Aircraft failure rate forecasting method based on Holt-Winters seasonal model. IEEE 2nd International Conference on Cloud Computing and Big Data Analysis (ICCCBDA). Chengdu.

Zacks, J., Levy, E., Tversky, B., & Schiano, D. (2002). Graphs in Print. In Diagrammatic Representation and Reasoning (pp. 187–206). London: Springer.

Zhu, G., Zheng, C., H. Hu W. G., & Shen, J. (2006). A Kind of Demand Forecasting Model Based on Holt-Winters Model and Customer-credit Evaluation Model. International Conference on Service Systems and Service Management. Troyes.

